# CLoSES: A platform for closed-loop intracranial stimulation in humans

**DOI:** 10.1101/2020.03.28.20040030

**Authors:** Rina Zelmann, Angelique C. Paulk, Ishita Basu, Anish Sarma, Ali Yousefi, Britni Crocker, Emad Eskandar, Ziv Williams, G. Rees Cosgrove, Daniel S. Weisholtz, Darin D. Dougherty, Wilson Truccolo, Alik S. Widge, Sydney S. Cash

**Author notes:** Corresponding author. Primary correspondence to: Rina Zelmann, Department of Neurology, Massachusetts General Hospital, 55 Fruit St. (Thier building, room 423), Boston, MA, USA 02114.

## Abstract

Targeted interrogation of brain networks through invasive brain stimulation has become an increasingly important research tool as well as a therapeutic modality. The majority of work with this emerging capability has been focused on open-loop approaches. Closed-loop techniques, however, could improve neuromodulatory therapies and research investigations by optimizing stimulation approaches using neurally informed, personalized targets. Specifically, closed-loop direct electrical stimulation tests in humans performed during semi-chronic electrode implantation in patients with refractory epilepsy could help deepen our understanding of basic research questions as well as the mechanisms and treatment solutions for many neuropsychiatric diseases.

However, implementing closed-loop systems is challenging. In particular, during intracranial epilepsy monitoring, electrodes are implanted exclusively for clinical reasons. Thus, detection and stimulation sites must be participant- and task-specific. In addition, the system must run in parallel with clinical systems, integrate seamlessly with existing setups, and ensure safety features. A robust, yet flexible platform is required to perform different tests in a single participant and to comply with clinical settings.

In order to investigate closed-loop stimulation for research and therapeutic use, we developed a <u>C</u>losed-<u>Lo</u>op <u>S</u>ystem for <u>E</u>lectrical <u>S</u>timulation (CLoSES) that computes neural features which are then used in a decision algorithm to trigger stimulation in near real-time. To summarize CLoSES, intracranial EEG signals are acquired, band-pass filtered, and local and network features are continuously computed. If target features are detected (e.g. above a preset threshold for certain duration), stimulation is triggered. An added benefit is the flexibility of CLoSES. Not only could the system trigger stimulation while detecting real-time neural features, but we incorporated a pipeline wherein we used an encoder/decoder model to estimate a hidden cognitive state from the neural features. Other features include randomly timed stimulation, which percentage of biomarker detections produce stimulation, and safety refractory periods.

CLoSES has been successfully used in twelve patients with implanted depth electrodes in the epilepsy monitoring unit during cognitive tasks, spindle detection during sleep, and epileptic activity detection. CLoSES provides a flexible platform to implement a variety of closed-loop experimental paradigms in humans. We anticipate that probing neural dynamics and interaction between brain states and stimulation responses with CLoSES will lead to novel insights into the mechanism of normal and pathological brain activity, the discovery and evaluation of potential electrographic biomarkers of neurological and psychiatric disorders, and the development and testing of patient-specific stimulation targets and control signals before implanting a therapeutic device.

## 1 Introduction

Neuromodulation with the use of direct electrical stimulation is a successful therapeutic approach for movement disorders (Lozano et al., 2019), pain (Levy et al., 2010), epilepsy (Morrell, 2006) and possibly for psychiatric disorders (Widge et al., 2018b). Although successful in some patients, current approaches are rudimentary, based on trial-and-error parameter selection. Success is measured on subjective appreciation of symptoms instead of a direct biomarker measuring the effect of stimulation in the brain. A wider variety of diseases could benefit from neuromodulatory approaches, but the effect of stimulation on human electrophysiology and the modulatory effect of stimulation on brain activity needs to be thoroughly investigated to inform the field on how to proceed (Lozano et al., 2019; Widge et al., 2017).

Most therapeutic stimulation devices provide continuous open-loop stimulation. Open-loop paradigms consist of short trains of direct electrical stimulation pulses that are delivered with an intermittent schedule. An exception is the RNS system (NeuroPace Inc, Mountain View, CA), that stimulates when a seizure is detected. It is the only closed-loop, or responsive, stimulator device approved by the FDA. Even in this case, it is not clear whether the effect is related to the closed-loop approach or simply to a long-term neuromodulatory effect of stimulation (Kokkinos et al., 2019). While the technology built into the RNS is indeed impressive it ultimately is designed for a low channel count system and a fixed and relatively simple set of feature analysis.

Closed-loop brain stimulation paradigms allow optimizing stimulation approaches using targeted, informed and personalized therapeutic strategies that could achieve better therapeutic results than open-loop stimulation and help understand underlying mechanisms. As stimulation can have very different effects with different brain states, brain regions, and pathological events, real time detection of these neural signatures during different brain states combined with stimulation can allow us to understand the ongoing networks. Detecting these brain states and applying the appropriate stimulation at the right time in the appropriate target could result in important therapeutic benefits including achieving better results, with fewer side effects, and longer battery life (Sun and Morrell, 2014; Tinkhauser et al., 2017). A flexible platform is needed to systematically perform diverse closed-loop tests in a clinical setting.

Most clinical research investigations in humans with semi-chronic implanted electrodes to localize the region responsible for seizure generation use open-loop direct electrical stimulation. For instance, during the intracranial pre-surgical workup of patients, electrical stimulation is helpful for functional mapping and to delineate the epileptic focus (Trébuchon and Chauvel, 2016). Clinicians try to elicit the patient’s typical aura or seizure or look for regions with low threshold to produce epileptiform after-discharges. As a research probe, single-pulse electrical stimulation (SPES; Valentín et al., 2002) or stimulation trains aim to understand neural mechanism and test network connectivity (Matsumoto et al., 2006; Trebaul et al., 2018) by quantifying the brain’s evoked response to stimulation, referred to as cortico-cortical evoked potentials (CCEPs; Matsumoto et al., 2004). Response to SPES varies depending on stimulation site and applied charge (Trebaul et al., 2018). Furthermore, modifying train stimulation parameters showed a nonlinear relation between stimulation response and frequency and a linear relation with current (Basu et al., 2019). These studies suggest intriguing characteristics of the brain’s response to stimulation and a possible effect of underlying rhythms. However, because stimulation occurs at random times, there is intrinsic low specificity when trying to study particular rhythms or cognitive states which results in imperfect, noisy measures and requires a large number of stimulations. Instead, we are proposing a closed-loop system which could have higher sensitivity and specificity to electrographic biomarkers.

Technological advances have fostered an increase in closed-loop neuroscience tests. Open source software such as Open Ephys (Siegle et al., 2017) and BCI2000 (Schalk et al., 2004) and hardware development of wireless implantable devices (Zhou et al., 2018) are greatly advancing the field. Substantial work in brain computer interfaces (BCI) in animals (reviewed in Fetz, 2015) and patients with implanted Utah electrodes in motor regions have achieved volitional motor control (Ajiboye et al., 2017; Bouton et al., 2016; Hochberg et al., 2006) and improved task performance (Katnani et al., 2016). Different feedback mechanisms have been used to close-the-loop (Wright et al., 2016). Feedback could be obtained from visual observation of movement (Hochberg et al., 2012), motion sensors (Espay et al., 2010), EMG activity (Kuiken et al., 2009; Nishimura et al., 2013), and, of particular interest to our work, neural activity (Ajiboye et al., 2017; Nishimura et al., 2013). Closing the loop to neural signals, by stimulating the brain following neuronal changes, creates a fully integrated feedback system but it is also very challenging (Krook-Magnuson et al., 2015). Most of the work in this realm focused on detecting single or multi-unit activity from neuronal ensembles (Collinger et al., 2013). However, intracranial local field potentials (LFP) measured with iEEG clinical macro-electrodes provide stable recordings for longer periods than single/multi-unit recordings (Flint et al., 2012). Therapeutically, LFP events detection reduces seizure occurrence (Morrell, 2006) and LFPs have been considered as control signals for closed-loop control in movement disorders (Tan et al., 2019; Tinkhauser et al., 2017) and pain (Shirvalkar 64 et al., 2018).

Closed-loop tests in humans could answer basic research questions and improve neuromodulatory therapies, but their implementation adds another layer of challenges (Caldwell et al., 2019). Developing systems that detect neural features and stimulate in real-time, deployed in the epilepsy monitoring unit (EMU) has technical, experimental, and design challenges. The technical challenges include: 1) the need to use continuous intracranial EEG (iEEG) activity which is recorded in a relatively noisy environment; 2) filtering at a variety of frequency bands; 3) implementing different control algorithms, such as power, coherence or cross-frequency coupling within the same framework; 4) ensuring low latency to detect and stimulate within the same physiological or pathological event; 5) guaranteeing precise processing times for seamless closed-loop operations; 6) implementing safety safeguards; and 7) ensuring reproducible tests. Experimental challenges during intracranial epilepsy monitoring, are due to electrodes implanted exclusively for clinical reasons and patients having different etiologies. There is little experimental control of specific channel locations, which vary across patients. Thus, detection and stimulation sites must be participant- and task-specific. Furthermore, the experimental paradigm involves hundreds of potential detection channels. This is challenging but could also be an advantage. With a flexible closed-loop system platform, different experiments could be performed in a single participant. In addition, the system would optimally run in parallel to clinical systems in different hospital rooms (e.g. epilepsy monitoring unit, intensive care unit, operating room) and integrate seamlessly with existing setups. From a design perspective, for the system to be used by investigators and clinicians with different level of technological expertise, it is important to: 1) have an easy to use intuitive interface; 2) easily switch across experimental paradigms, each with task and participant specific channels, features, or state models; 3) have real-time visualization of ongoing iEEG activity, features, thresholds, detections and stimulation to allow confirmation of accurate experimental setup and manual parameter adjustment in real-time; 4) allow investigators to visualize stimulation-locked average iEEG activity to ensure that response to stimulation can be obtained; and 5) allow for offline replay of the data to personalize parameters and train algorithms.

To overcome these challenges, we introduce a general-purpose platform for <u>C</u>losed-<u>Lo</u>op <u>S</u>ystem for direct <u>E</u>lectrical intracranial <u>S</u>timulation (CLoSES) based on real-time decoding of intracranial brain signals, together with a pipeline for closed-loop tests in humans. CLoSES fills a critical niche between animal investigations where highly flexible equipment can be used without regulatory constraints and implantable therapeutic devices in humans where the degrees of freedom are very limited. CLoSES provides a new tool to precisely probe the human brain, solving the need of a general framework tailored for the unique space of iEEG recordings in humans.

In the methods section, we describe CLoSES’ software and hardware design, with a pipeline for closed-loop tests. In the results, we present four applications where we implemented and utilized the system in humans in the EMU and discuss performance capability. Finally, in the discussion, we summarize the system’s configuration, strengths and weaknesses and discuss its place in burgeoning avenues of basic and clinical neuroscience research.

## 2 Materials and Methods

We developed a <u>C</u>losed-<u>Lo</u>op <u>S</u>ystem for <u>E</u>lectrical <u>S</u>timulation (CLoSES) that computes neural features which are, in turn, used in a decision algorithm to trigger stimulation in near real-time. To ensure fixed processing times, CLoSES runs on a dedicated Simulink Real-Time (MathWorks) computer. This allows iEEG acquisition and processing (filter, compute features, detect) every millisecond. A separate computer runs the graphical user interface (GUI) for configuration and real-time visualization. CLoSES seamlessly runs in parallel with 24/7 clinical and research acquisition systems, avoiding intereference with clinical equipement in the EMU. CLoSES accepts ancillary inputs that can be used to trigger threhold updates, feature calculation, and stimulation time. Off-line replay of previously acquired datasets allows parameter optimization and feature analysis. This is useful to obtain patient and task-specific features, parameters, and models. Implemented safety features are of importance to ensure limitted stimulation delivery as CLoSES is utilized in the hospital setting.

Figure 1 shows the general hardware diagram of the complete rack-mounted system. It consists of the iEEG acquisition hardware; a *presentation* computer to run experimental tasks; a *stimulation* system that sends a stimulation pulse or train upon receiving a stimulation trigger; and the closed-loop system, CLoSES, which is described in detail in this manuscript. CLoSES is composed of a Windows *host* computer and a dedicated *target* computer with Simulink Real-Time kernel. The host computer allows GUI visualization, data saving and configuration, while the target computer ensures fixed time processing for real-time computation.

**Figure 1.**
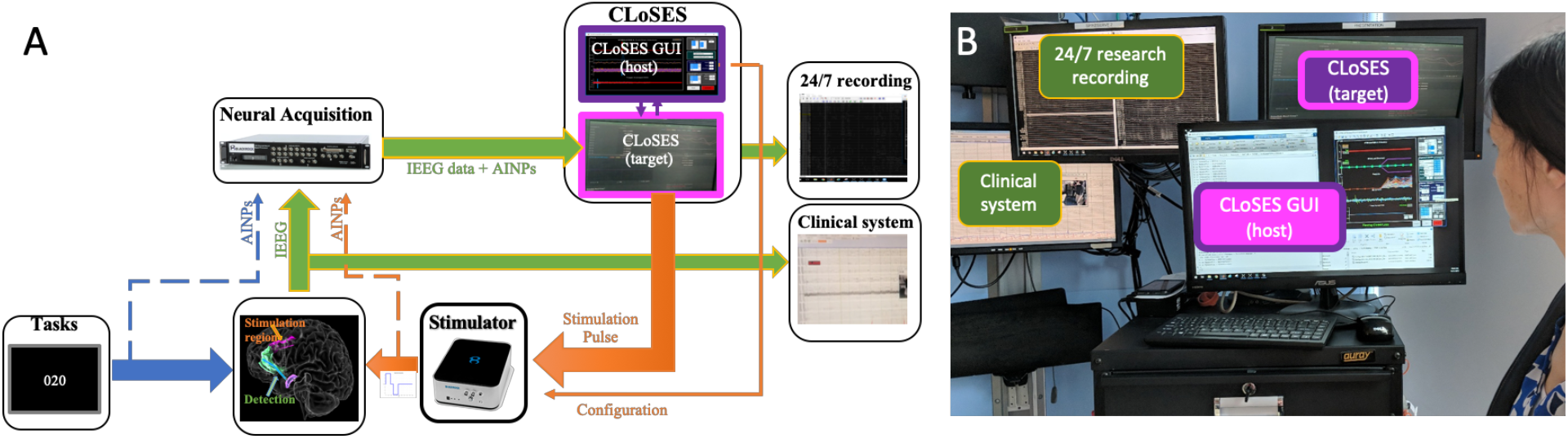
A) Schematic of closed-loop experiments. B) Example of CLoSES in the EMU. The closed-loop system consists of intracranial EEG acquisition, a stimulation system, a computer to present cognitive tasks, and CLoSES. CLoSES is composed of a GUI for visualization, data saving, and configuration (*host*, that runs under Windows) and a dedicated target computer with Simulink Real-Time kernel to ensure fixed time processing for real-time computation. The iEEG data are acquired in parallel by the clinical system, the 24/7 research acquisition equipment, and CLoSES. Green arrows indicate iEEG data, *blue* arrows are related to task, *orange* arrows are related to stimulation. *Dotted* arrows indicate ancillary inputs (AINPs).

Two modes of operation were implemented: R**eal-**T**ime CLoSES (CLoSES-RT)**, to stimulate in real-time following continuous neural feature detection and **CLoSES with Neural** S**tate** E**stimate Encoder/Decoder** M**odel (CLoSES-SEM)**, to stimulate following decoded hidden states during cognitive tests.

The complete source code for these CLoSES variants are available on GitHub (https://github.com/Center-For-Neurotechnology, CLoSES-SEM and CLoSES-RT repositories), along with instructions for compilation and deployment. CLoSES is licensed under the BSD 3-Clause Clear License. Depending on the specific hardware involved, specific manufacturer software development kits (SDKs) may also be needed. For instance, the examples here used datagram decryption and stimulator control libraries from Blackrock Microsystems (Salt Lake City, UT).

### 2.1 CLoSES-RT: Stimulate in near real-time following neural feature detection

In CLoSES-RT, iEEG brain signals are acquired, re-referenced if needed, band pass filtered, and neural features continuously computed. Implemented features are: coherence across channels and multi-band power per channel. These features are computed at each time step as an average over a time interval. As in a particular previous application (Sarma et al., 2016), we implement a dual-threshold control algorithm: if features are above (below) the upper (lower) threshold for a certain duration, stimulation is triggered. Thresholds can be manually adjusted or dynamically updated as detailed in section *Threshold Update*. Stimulation can occur following detection, after a fixed delay, or at the time of an external input. To study the effect of stimulation at the time of a spontaneous event, it is necessary to compare detected and stimulated events to interleaved random stimulation and to detected events that were not stimulated. The rate of random stimulation and the percentage of detections that produce stimulation can be specified. The fixed step size can also be modified (default is 1ms). Safety refractory periods prevent semi-continuous stimulation and an initial block-out period ensures stimulation occurs only under stable software and hardware conditions. Offline replay of data allows parameter optimization and channel selection. Figure 2.A shows the block diagram of CLoSES-RT and Figure 2.B an example of GUI visualization while CLoSES-RT is running in the EMU.

**Figure 2.**
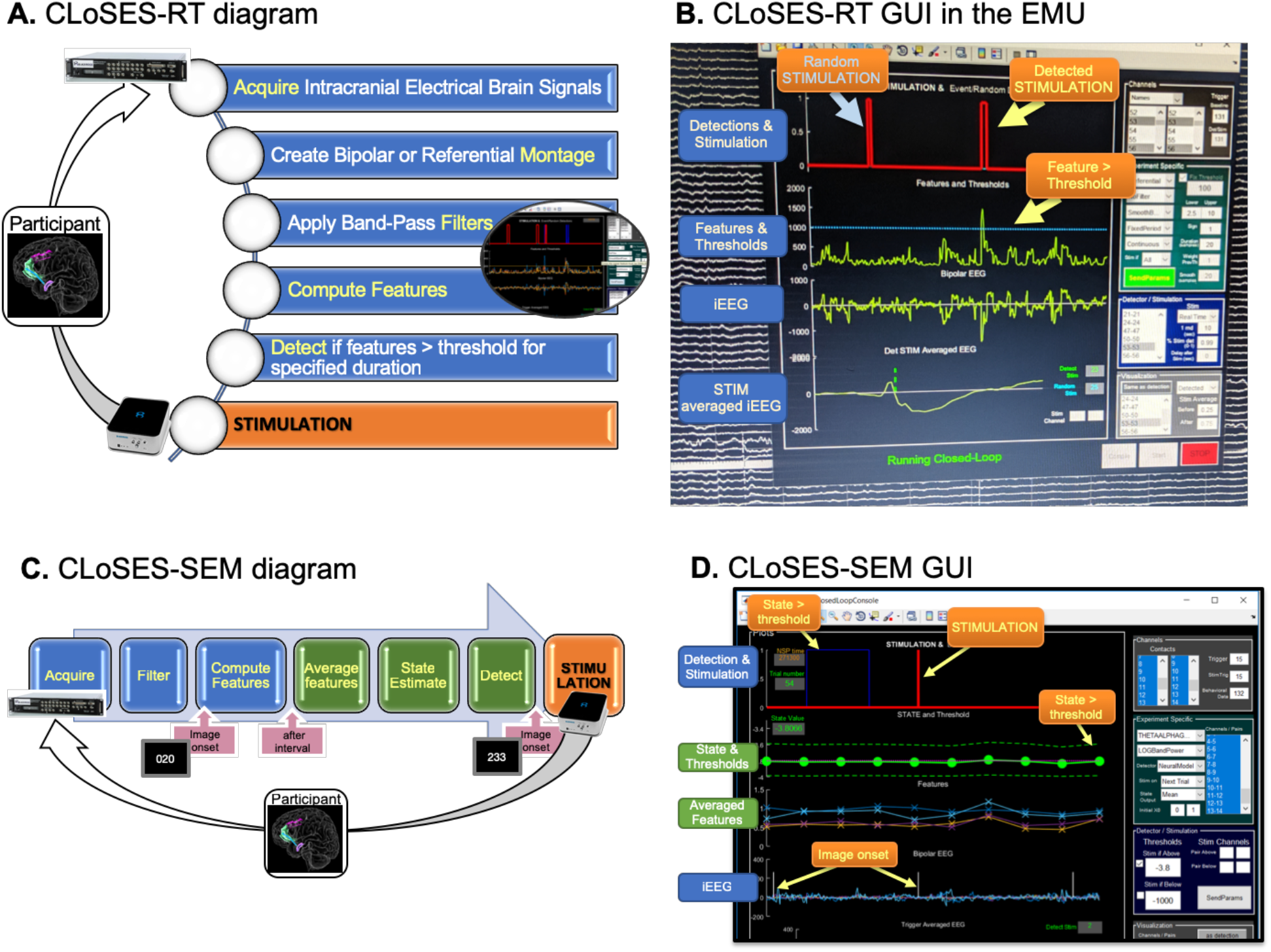
Diagram and GUI of CLoSES-RT and CLoSES-SEM. A) Diagram of CLoSES-RT. Neuronal iEEG signals are acquired, re-reference, band-pass filtered, and features computed in real-time. In the example, when power in detection channel is above threshold for certain time (20ms), a stimulation pulse is sent. The detection process is repeated every millisecond. B) CLoSES-RT GUI showing ongoing neural activity during an experiment in the EMU. Intracranial EEG activity, features, threshold, detections, and stimulation of selected channels are updated every 100ms. When stimulation occurs, the averaged iEEG plot is also updated. Most common configuration parameters can be selected in the right panel. C) Diagram of CLoSES-SEM with neural decoder model. Intracranial EEG signals are continuously acquired and band-pass filtered. Following image onset trigger, features are continuously computed. After a specified interval (default: 2 seconds), features are averaged in epochs and provided as input to the neural decoder model to estimate cognitive state. In the example, when mean state estimate is above threshold a stimulation train is sent at the time of the following image onset. D) CLoSES-SEM GUI showing ongoing visualization. IEEG, and stimulation are updated every 100ms. Averaged feature, state and threshold are updated every trial. Most common configuration parameters can be selected in the right panel. Blue boxes indicate continuous steps, green boxes indicate once per trial steps, boxes with arrows indicate external events.

### 2.2 CLoSES-SEM: Stimulate following decoded cognitive state

In the case of CLoSES-SEM, a state-space model (Yousefi et al., 2019a) estimates the brain’s “state” from online recorded iEEG during the performance of cognitive tasks. That is, at each trial, features (power, log power or coherence) are continuously computed as in CLoSES-RT, but are then averaged for specific task related epochs, and are passed to a model to compute a cognitive hidden state. If decoded state exceeds (is below) threshold, stimulation is triggered. The model could range from a simple weighted combination of averaged features to an encoder/decoder model based on the assumption that it is possible to decode the latent-variable, or unobserved cognitive state of the brain, from neuronal activity (Yousefi et al., 2019a). The model can be optimized offline on non-stimulation and/or open-loop stimulation task data. Figure 2.C shows the block diagram of CLoSES-SEM and GUI visualization.

Table 1 summarizes the common steps and differences between CLoSES-RT and CLoSES-SEM default mode of operation.

**TABLE 1.** Comparison of operation for CLoSES-RT and CLoSES-SEM approaches.

| Common Steps |  |  |
| --- | --- | --- |
| Acquisition | Continuously acquire intracranial electrical brain signals from NSPs or Acquisition boards. |  |
| Montage | Create Bipolar or Referential Montage. |  |
| Filters | Band-Pass Filters. |  |
| Features | Compute Features (power, coherence). |  |
|  | CLoSES-RT | CLoSES-SEM |
| Control Signal | Compute Features continuously. | Compute Features per trial, averaged over time epochs, and a neural encoder/decoder model estimates a hidden state. |
| Detection | If features > threshold for certain duration. | If decoded state > threshold. |
| When to send stimulation | Trigger stimulation in real-time. | Stimulation occurs on the following trial. |
| Comparison | Interleaved Random stimulation. | Stimulation not linked to state variable. |
| Processing | Fixed steps = 1ms. | Fixed steps = 10ms. |
| Safety | 2 sec refractory period. | Stimulate only N out of every M trials. |

### 2.3 Graphical User Interface

We developed a GUI for parameter configuration and real-time visualization. For CLoSES-RT, iEEG, features, thresholds, detection, and stimulation signals are updated every 100ms (Figure 2.B). For CLoSES-SEM, iEEG, detection, and stimulation data are updated every 100ms. Features, thresholds, and state estimate (mean and boundaries) are updated every trial (Figure 2.D). In addition, to help visualize the response to stimulation, both GUIs allow real-time update of stimulation averaged iEEG under different conditions (e.g. random or detected events). In this way, the effect of stimulation on the brain can be visualized as the test progresses. An advantage of using a MATLAB environment is that all figure functionalities are available. For instance, when zooming in, all continuous plots and all trial by trial plots change together. When clicking on a plot, the y axis scale is adapted for best fit. This allows quickly checking something in detail on the ongoing signal and coming back to visualizing five seconds and 10 trials simultaneously.

The right panel allows parameter configuration of the most common parameters (Figure 2.B & 2.D). For instance, the GUI allows channel, montage, frequency bands, feature type, and model selection. When to stimulate, threshold amplitude, detection duration, or how to update thresholds can also be configured. Some parameters can only be selected at the beginning of the test, while others can be modified in real-time. Parameters that can be updated in real time are those that can change with brain state, or that may need to be adjusted during the experiment. The most common are: threshold (either the value or a multiplier over a computed baseline), minimum duration of detection, and rate of random stimulation. Parameters that can only be modified at the beginning of the experiment include those that define the path of the Simulink system (e.g. frequency bands, feature), since a new compiled file is sent to the target computer if these parameters are changed, and parameters that are in general the same for different participants (e.g. refractory period, smoothing window). In addition, all parameters can be specified through patient and task specific configuration files that loaded as initial values in the GUI. For a complete list of parameters see online manual on GitHub release. Supplementary videos V1 and V2 show examples of GUI operation during offline replay data.

#### 2.3.1 Configuration Files

In addition to parameter selection through the GUI, it is possible to use participant and task specific configuration files. This includes presetting of GUI parameters, configuration of less common parameters not visible on the GUI, and stimulation parameters. Pre-loading the configuration prevents human operator errors, especially with channel selection. It allows rapid switching between experimental paradigms with different channels, features, and models. Configuration files can also be used to replicate setting from another test, by loading saved parameters. Participant and task specific configuration files are usually created after analyzing replay of data acquired during non-stimulation (training) days (see also *Replay Simulations vs. Real-Time data acquisition* section).

### 2.4 Intracranial EEG Data Acquisition

CLoSES can flexibly acquire data from multiple common human electrophysiology systems. Two acquisition inputs have been developed: UDP acquisition from BlackRock neural signal processors (NSPs) and National Instruments analog input card, connected to a Plexon (Dallas, TX) system. The modular approach allows other input blocks to be added with minimal changes to the original design. All the examples in this paper were acquired in the EMU using BlackRock NSPs. In this configuration, the intracranial EEG signal is split from the clinical Natus system (Natus Medical Inc., Pleasanton, CA) and transmitted through an Ethernet connection as UDP packets from the NSPs to both CLoSES and a computer running BlackRock central software for 24/7 recording. The iEEG data are acquired by capturing and parsing the UDP packets. Thus, CLoSES runs in parallel to clinical and research recording systems. In our setup, given the large number of electrodes normally implanted, and since each NSP can record up to 128 EEG channels, two NSPs are usually used simultaneously.

When running a behavioral task, event markers (triggers) are sent from the computer presenting (presentation computer) the task and are connected to an ancillary analog input (AINP) on the NSP, that is acquired together with the iEEG data. This input can indicate when to start computing features, when to update thresholds, and when to stimulate. For instance, during cognitive tasks, features can be computed for each trial from the beginning of a trial (the input is referred to as image onset, when image appears) and stimulation could occur at the time of the following trial (following image onset). Outputs from CLoSES, indicating stimulation and detections, are also connected to AINPs. This is useful for instance to avoid detections immediately following stimulation, to implement safety constrains, and to compute stimulation-locked averages. If another computer is calculating an important feature, e.g. a performance variable estimated directly from behavior (Paulk et al., *submitted*), this can be directly connected to another AINP for simultaneous real-time visualization of neuronal and behavioral state estimates.

### 2.5 Channel Selection and Montage Generation

IEEG is acquired referenced to one of the leads (in our institutions this is often Cz on the scalp but can, of course, be any channel). The acquired iEEG signal could be used in referential montage or re-referenced to bipolar montage. Bipolar montage is usually created from consecutive contacts within an electrode shaft, but different contact combinations could be specified on configuration file or selected on the GUI. This can be useful to detect around stimulation contacts or if a contact is broken. Similarly, the AINPs channels where stimulation return, behavioral data, or image onset trigger are connected could be specified. Triggers are detected as rising edge.

A bipolar montage is useful to cancel noise and focus on local characteristics recorded by iEEG. A referential montage is particularly useful when stimulating and recording from a small region. As an example, during closed loop to IIDs in the hippocampus, where only three contacts of the shaft are within the region of interest, we can use one contact for detection and the consecutive two contacts for bipolar stimulation.

### 2.6 Filters

The following traditional frequency bands are implemented with IIR Butterworth filters: Theta (3-9Hz; order 24), Alpha (8-15Hz; order 24), Spindles (11-15Hz; order 2), Beta (15-30Hz; order 26), LowGamma (30-55Hz; order 26), HighGamma (65-110Hz; order 32), and Ripple (80-200Hz; order 32). A refractory period following stimulation reduces the contribution of stimulation artifacts. Multi-band filter banks are easily implemented as a parallel combination of filters.

### 2.7 Features

Currently the following features are implemented: *smooth power, log power*, and *coherence*. Given the modularity of the system, new features can be easily added or combined. Features are computed continuously at every step, following time domain band-pass filtering of the iEEG. Depending on the feature, the calculation is bounded (e.g. 0-1 for coherence) or not. In CLoSES-SEM, features are then averaged per specified epoch within the window of analysis (e.g. there could be four 500ms epochs in the two seconds following image onset).

#### 2.7.1 Power Implementation

The *smooth power* feature is obtained as the running average of the root mean square of the band-pass filtered signal, over a specified time interval. This averaging interval means that at each time point the smooth power value is the average power of the previous samples. Default is 50ms, but to obtain short latencies, a shorter interval could be specified. For instance, for IID detection we used 10ms. Multiple frequency bands can be computed in parallel. In CLoSES-SEM, logarithm is applied to obtain a normally distributed input to the decoder model. As only a few features per trial are need, the average *log power* per epoch (default: two epochs of 500 ms each) is computed. Thus, multi-band *log power* is computed continuously, then averaged per specified epochs, and used as input features for the decoder model.

#### 2.7.2 Coherence Implementation

The magnitude square coherence function from MATLAB (*mscoherence*.*m*), is not compatible with Simulink Real-Time. In addition, it takes unnecessary time as it computes power three times. Thus, we wrote a Simulink Real-Time compatible coherence function. Our custom coherence function is based on DFT with chirp-z transform, contains pre-initializing variables, fix number of FFT frequencies. It uses a matrix form and computes only the necessary power spectra. In brief, the algorithm is the following:

1. From the pairs of interest obtain the individual channels.
2. Compute auto-spectrum (S_xx_) for each channel and *k*^*th*^ *data segment*
  ○ For each segment to create the periodogram:
    ▪ Compute the Discrete Fourier Transform (DFT). Use the chirp-z transform since it is the only method that could be adapted to Simulink Real-time

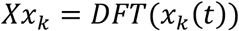
    ▪ Compute the power by multiplying to its conjugate

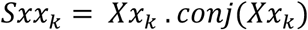
    ▪ Keep Sxx_k_ and Xx_k_ for each segment.
  ○ Average *K* segments to obtain periodogram (default K=4)

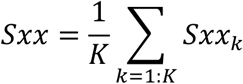
3. Compute cross-spectrum (S_xy_) for channels of each pair:
  ○ Since we already computed all the individual DFTs for each segment, we can simply multiply the Xx_k_ of the two channels of a pair (denoted Xx_k_ and Yy_k_) to obtain the cross-spectrum

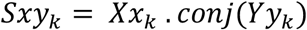
  ○ The cross-periodograms are obtained by averaging the *K* segments

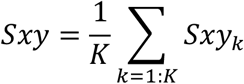
4. Compute Coherence for all pairs

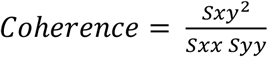

Coherence is computed continuously. At each time point the coherence value is computed buffering previous samples (default 500ms). When computing coherence there are always trade-offs to consider. Because most of our applications to date use low frequency bands, we use as default long time buffer (500ms) and only 4 bins per frequency band for the periodogram with 50% overlap. This implementation is computationally efficient. During replay it takes about 3 hrs to run 100 channels (4950 features). This enables the use of CLoSES to replay participant-specific data offline for optimal feature selection and decoder model training. In CLoSES-SEM coherence is then averaged per epoch of interest (default: 2 epochs of 500ms each) and used as input to the decoder model detailed below.

### 2.8 Decoder Model Implementation

Averaged features are the input to the neural decoder model. Decoder model complexity range from a simple weighted combination of features to a neural encoder-decoder model created with COMPASS software package (Yousefi et al., 2019b). To select optimal features and train the model, features of a large number of channels are computed offline, using replay of datasets acquired during prior test runs with the participant. During the closed-loop tests, one-step decoding is based on the features calculated as explained above, using the model parameters and features selected from this offline training.

The online decoder is essentially the same as the published offline one (Yousefi et al., 2019a). Re-organizing data structures and algorithms and using constants instead of variables made it compatible with Simulink Real-Time. Online decoder implementation is the same for the two examples presented, even though the encoder models are different and one approach uses power (Basu et al., *submitted*) while the other uses coherence as the underlying features (Paulk et al., *submitted*). Given the CLoSES modularity, new models could be easily incorporated.

### 2.9 Detectors

In CLoSES-RT, detection occurs if a threshold is crossed in the selected direction/s for a specified number of samples. The stimulation pulse can be sent in real-time or with a predefined, fixed delay. In CLoSES-SEM, detection occurs based on the mean, upper and/or lower confidence bounds of the estimated state. A stimulation pulse or train is sent if the selected output from the decoder model is above the upper threshold or below the lower threshold. The pulse can be sent in real time or in response to a task event during the following trial. To achieve bidirectional control, different pairs of stimulation channels can be configured in the GUI. For instance, if upper state bound is above the upper threshold, stimulation is delivered on bipolar channel one, if lower state estimate bound is below the lower threshold, stimulation is delivered on bipolar channel two.

#### 2.9.1 Threshold Update

Thresholds can be updated in a variety of ways depending on the application (Table 2). 1) The simplest option is to have fixed thresholds and modify them in real time using the GUI. 2) It is also possible to compute thresholds based on triggers from ancillary external inputs, for instance to compute per-trial thresholds as an N-fold increase over a baseline period. 3) Thresholds could be updated periodically (e.g. every minute), to account for loss of stationarity in the signal. 4) Thresholds could be updated after N stimulations, to consider the long-term effect of electrical stimulation on the network. 5) Combining these options is also possible, as thresholds could also be updated after N stimulations or after X seconds without any detection.

**TABLE 2.**
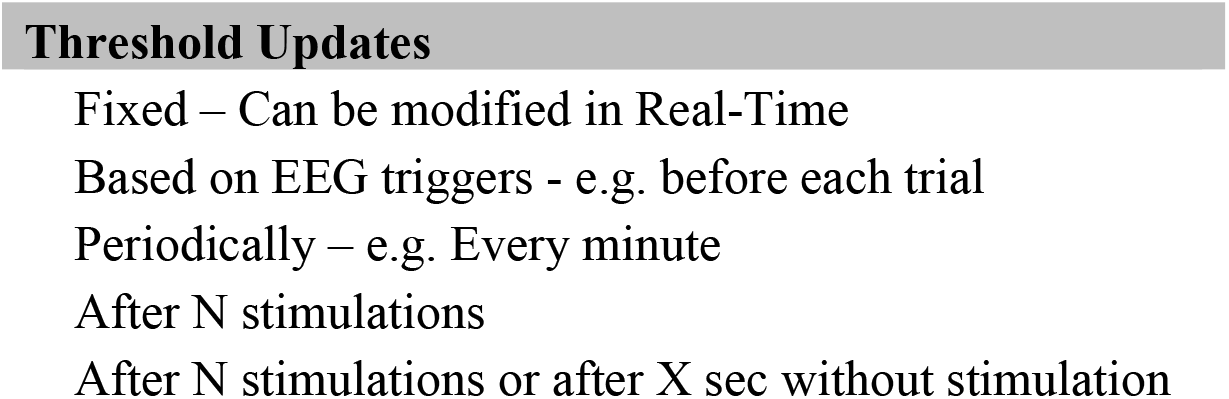
Threshold update implementation options.

### 2.10 Stimulation options

A stimulation pulse triggered via TTL is sent when detection occurs, after a fix delay, or at a trigger from an ancillary external input following detection. The configuration of single pulse or pulse train frequency and amplitude are set at the beginning of each test. In the case of single site stimulation, the pair of channels where stimulation is delivered is also configured once before starting the test. In the case of multisite stimulation for bidirectional control, stimulation can occur at two pre-configured sites. As explained in section *Detectors*, the pair of channels to stimulate is selected depending on which detection (higher than upper threshold or smaller than lower threshold) occurs. A set of custom made functions based on MATLAB API for BlackRock Cerestim (https://github.com/Center-For-Neurotechnology/CereLAB) allow us to connect to the stimulator and change its configuration. This action takes around 100ms. Bi-directional control was important during cognitive tests, in which stimulation channel is changed after detection and on the following trial a TTL pulse to trigger stimulation is sent. Because the Cerestim API cannot be compiled in Simulink Real-Time, we implemented stimulation channel configuration in the host computer. In all cases, a parallel port channel sends a TTL pulse to trigger stimulation.

For safety reasons, a ‘block out’ period at the beginning of the test and a refractory period (default two seconds) in which stimulation cannot occur can be configured. In addition, only N out of M trials can be stimulated. For example, with N=5 and M=10: if trials #2 to #6 resulted in stimulation, the next trial in which stimulation could occur would be trial #12. Stimulation characteristics are summarized in table 3.

**TABLE 3.**
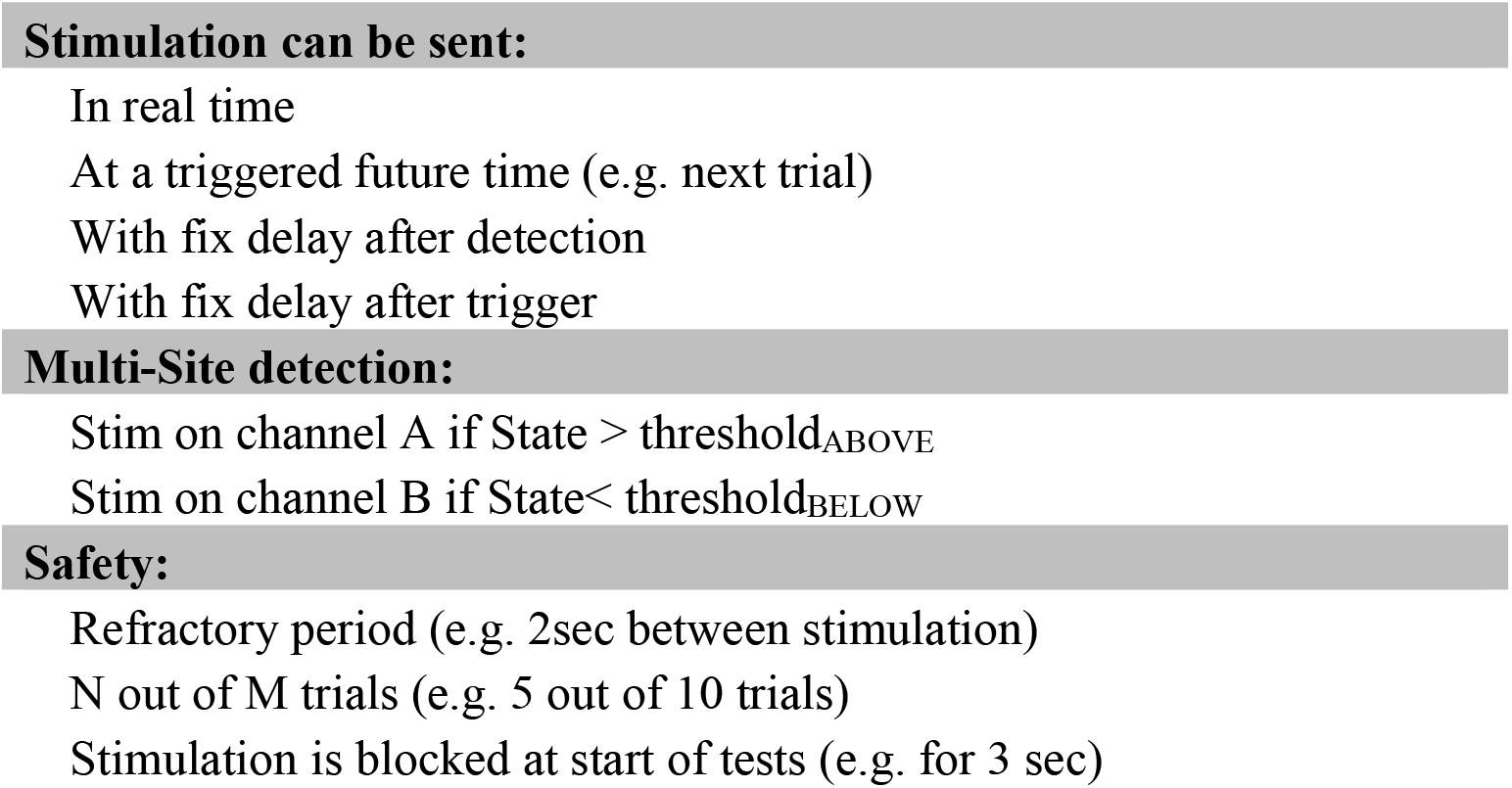
Stimulation options.

In terms of hardware, Cerestim (Blackrock Microsystems, Salt Lake City, UT) and A-M Systems Model 3800 (A-M Systems, Sequim, WA) stimulation systems interfaces were implemented. In both cases, configuration is done on a Windows computer on the stimulator default software or through provided API. The stimulator is configured to “wait for triggers”. When detection occurs, CLoSES sends a TTL pulse to trigger stimulation. Thus, any stimulator could be used with no need of reconfiguration or programming in CLoSES.

### 2.11 General Characteristics

#### 2.11.1 Fixed time implementation

In CLoSES-RT, montage configuration, band-pass filtering, feature calculation, and detection occur at a default fixed step of 1ms. This is an important characteristic for a closed-loop system that operates in near real-time. In CLoSES-SEM, during cognitive tests organized in trials, the default is to detect a neural signature during one trial and decide whether to stimulate or not at the start of following trial. Thus, we do not actually need that speed for the cognitive tests. Power calculation was within the 1ms step, but computing coherence (even with the fast new implementation) for more than 5 channels/ 10 pairs takes longer than 1ms. We therefore decided to increase the step size to 10ms. The system still acquires at 2000Hz, but processes every 10ms. This is below most BCI implementations (Wilson et al., 2010). Executing a step of the decoder model takes longer, but since this step is computed only once per trial, at the end of the continuous feature calculation, the system has time to recover until the following trial (See details in *Performance analysis*).

### 2.12 Replay Simulations vs. Real-Time data acquisition

All processing and control algorithms are the same for offline replay and real-time operation. Replaying of previously acquired datasets is useful to select features, for model training, to analyze parameters, and for visualization. In addition, it allows comparison of algorithms and detection strategies. For replay to be useful, it must produce the same features and make the same decisions regarding when to stimulate as in the real-time system. Thanks to our modular system design, we can ensure that the exact same algorithms are applied. Indeed, only the input block is different when doing real-time acquisition and processing or replaying previously acquired data. Depending on the acquisition system, acquisition delay must be considered when comparing offline replayed and acquired data (for our setup see Performance section). Importantly, replay simulations can be run on any Windows computer (Simulink Real-time requires a dedicated computer or Windows operating system). As this is a replay, there is no limitation on processing time per step. It is possible to run any number of channels and features, which is particularly important to train the encoder model with features generated exactly in the same way as will be obtained during the closed-loop tests.

### 2.13 Performance analysis

In CLoSES-RT, detection was based on neural features directly and stimulation could be selected to occur at following AINP input, with delay, or immediately following detection. For the later, it was important to ensure near real-time processing. We therefore configured our system to have a 1ms time step. We quantified the real duration of each step to ensure that we were within the specified fixed value. Hardware stimulator delay was 2µs (Blackrock, personal communication) and will be ignored in subsequent analysis. When using Blackrock acquisition, UDP latency was 6-7ms. Within CLoSES-RT, total latency from start of an event to stimulation is the composite of filter order, feature buffering time, smoothing windows, and detection interval, times the fixed-step duration.

On CLoSES-SEM, we can stimulate in real-time following detection or when an ancillary input indicates a new trial. Since the latest is the most common implementation, the real-time constraint can be relaxed. In this case, features are only computed from image onset up to one or two seconds and averaged. Then, there is about one second until the stimulation pulse is sent. Thus, it was important to be within the fixed step for the continuous steps (acquisition, filtering and power calculation), but we could relax this constraint for blocks that are executed only once per trial. In addition, we could implement higher order filters. We configured our system to have a 10ms fixed step, removed the fixed step constraint in the decoder model block, and added rate transitions and buffers to ensure a robust implementation.

We evaluated whether processing time increased linearly with number of channels and features. We computed histograms of real duration for each step and assessed whether they were within the specified fix-step. We tested CLoSES-RT for 3, 5, 6, and 10 channels. We tested CLoSES-SEM for 10, 20 and 50 channels for power calculation (30, 60, and 150 features) and for 5, 10 and 20 channels for coherence (10, 45, and 190 pairs). This is more channels and features than would be used in practice in an implantable system, as usually 5-10 channels are selected and only a subset of features is used in the models (Widge et al., 2017, Basu et al., *submitted*).

### 2.14 Participants

Closed-loop tests were performed in twelve patients with semi-chronic electrodes implanted exclusively for clinical reasons at the Massachusetts General Hospital (MGH) or Brigham and Women’s Hospital (BWH). Depth electrodes (Ad-tech Medical, Racine WI, USA, or PMT, Chanhassen, MN, USA) with diameters of 0.8-1.0mm and consisting of 8-16 platinum/iridium-contacts 1-2.4mm long were stereotactically placed for seizure localization. iEEG was acquired with a Blackrock system at 2 kHz sampling rate (Blackrock Microsystems, Salt Lake City, UT, USA) referenced to a scalp contact.

All patients voluntarily participated after fully informed consent according to NIH and Army Human Research Protection Office (HRPO) guidelines as monitored by Partners Institutional Review Board (IRB). Participants were informed that participation in the tests would not alter their clinical treatment in any way, and that they could withdraw at any time without jeopardizing their clinical care.

In the examples presented in this manuscript, CLoSES-RT was used to detect and stimulate during epileptic IIDs and sleep spindles. CLoSES-SEM was used during a Multi-Source Interference Task (MSIT) to study the effect of stimulation on cognitive flexibility and during an Emotion Conflict Resolution (ECR) task to study the effect of stimulation on the regulation of emotion. Table 4 shows the type of tests performed with each participant.

**Table 4.** Type and number of tests per subject.

| Participant | CLOSES-SEM |  | CLOSES-RT |  |
| --- | --- | --- | --- | --- |
|  | MSIT | ECR | IIDs | spindles |
| S1 |  | 1 |  |  |
| S2 |  |  | 1 |  |
| S3 |  | 1 | 1 |  |
| S4 | 1 |  | 1 | 1 |
| S5 | 1 | 1 | 1 |  |
| S6 |  |  | 1 | 1 |
| S7 |  |  |  | 1 |
| S8 | 1 |  |  | 1 |
| S9 |  |  | 1 |  |
| S10 |  |  | 1 |  |
| S11 |  |  | 1 | 1 |
| S12 |  |  | 1 | 1 |
| **TOTAL** | **3** | **3** | **9** | **6** |

#### 2.14.1 Stimulation Parameters

Stimulation was delivered with CereStim R96 stimulator (Blackrock Microsystems, Salt Lake City, UT). SPES or train pulses can be configured directly in the stimulator software or using the API from CLoSES. In the examples presented here, IID and spindle tests used SPES. MSIT and ECR tests used trains of 130 Hz and 160Hz respectively. Individual biphasic pulses were 90μs long each with 53μs inter-stimulus interval. Current intensity was decided during open-loop safety testing in the stimulating channels, resulting in 2-7 mA.

### 2.15 Pipeline for CLoSES tests in humans

As important as developing a platform for closed-loop was to implement a clear pipeline for closed-loop tests with human participants in the EMU. In these cases, patients typically stay for 7-15 days to record enough seizures via intracranial leads. Research stimulation tests were performed after the clinical team obtained all the relevant clinical information and following restoration of antiepileptic medication (usually the day before electrode explantation). Safety testing of possible stimulation channels, consisting on the same stimulation parameters (SPES or trains) at intensities up to 7mA, was performed before CLoSES tests. An experienced epileptologist reviewed the iEEG online to ensure that no after-discharges were elicited. After each stimulation block, the participant was asked if s/he felt anything. Only if the iEEG did not indicate aberrant neural activity and the participant did not experience any subtle sensation, the site was eligible for CLoSES-driven stimulation.

The following pipeline was used for CLoSES-RT tests. Simulation data for replay was created from ongoing recordings during the same state (awake or asleep) as intended for the closed-loop test. Physiological events of interest (e.g. IIDs or spindles) were visually identified. Using offline replay of this simulated data channels were selected and parameters were optimized to detect exclusively the events of interest. Configuration files were created with the participant and test specific configuration. During the closed-loop test in the EMU: 1) Participant and task specific channels and parameters were loaded from configuration files. 2) CLoSES-RT was run without stimulation to confirm that channels and parameters could successfully detect the events of interest. If needed, parameters were modified. 3) To obtain interleaved stimulation outside the events, random stimulation frequency was selected. 4) Stimulator was turned on and CLoSES-RT tests started. During the tests some parameters such as threshold could be adjusted if needed. 5) Data are saved in 24/7 research computer, CLoSES host computer, and as backup in CLoSES target computer.

The following pipeline was used for CLoSES-SEM tests. Participants performed at least a block (64 trials) of the cognitive task without stimulation. Simulation datasets were created from these data and replayed offline. All possible combinations of reasonable features were processed based on replay of these simulations. Model optimization, including feature pruning, created a participant specific model (Paulk et al., *submitted*, Basu et al., *submitted*), with optimal parameters and selection of a subset of channels and features. To increase variability and further decrease feature dimensionality, an open-loop stimulation training step could be performed. In this case, during randomly selected trials of the cognitive task a stimulation train was delivered. During the closed-loop test: 1) Subject specific and task specific model, parameters, and channels, were loaded from configuration files. 2) The participant performed a block of the task without stimulation to obtain a baseline state, with CLoSES-SEM running in the background to check state model estimate. 3) Stimulator was turned on and closed-loop tests started, without the participant feeling anything. The participant performed 2-3 blocks of the experiment. Some parameters such as threshold could be adjusted as needed. 4) Participant performed one more block without stimulation as control. 5) Data were saved in 24/7 research computer, CLoSES host computer, and CLoSES target computer.

## 3 Results: Examples of application

In this section we illustrate the use of CLoSES by providing example of applications of CLoSES-RT and CLoSES-SEM. CLoSES has been tested in twelve patients with implanted depth electrodes in the EMU. We first provide examples of CLoSES-RT during IIDs detection (N=9) and sleep spindle detection (N=6). In these examples, iEEG was acquired at 2000Hz, processing time step was set to 1ms, and stimulation was sent in real-time. We then present examples of CLoSES-SEM during the MSIT cognitive flexibility task (N=3) and ECR emotion regulation task (N=3). In the these examples, iEEG was acquired at 2000Hz, processing time step was set to 10ms and stimulation was sent on the following trial. These examples illustrate the flexibility of CLoSES in various experimental paradigms. In addition, we present results of performance analysis.

### 3.1 Application #1: CLoSES-RT detecting interictal discharges in epileptic tissue

The goal was to examine the association between IID events and brain excitability by delivering stimulation during detected IIDs (Sarma et al, 2016). We assessed the effects of SPES stimulation on IIDs and the effect of IIDs on CCEPs following SPES, across different brain areas. These large amplitude short-duration events were detected with the following parameters in CLoSES-RT: unfiltered iEEG, smooth power (5-10ms window average), detection occurred if power was above threshold for a duration of 10-25ms. When an IID was detected, SPES was delivered to an adjacent bipolar channel in real-time to stimulate during the large epileptic spike or with a fixed delay to stimulate during the slow wave. The delay was estimated for each patient using offline replay of previously acquired data. When detecting IIDs, only one or a few channels are of interest. Figure 3 shows examples of detected and stimulated IIDs, random stimulation, and detected but not stimulated IIDs. In the first example, IIDs were detected in referential montage in the deepest contact of the right posterior hippocampus and stimulation was delivered to the nearest bipolar channel. This ensured that detection and stimulation occurred within the same anatomical structure. Increased amplitude can be observed just before detection during detected and stimulated IIDs compared to random stimulation. After stimulation, iEEG for the detected IIDs is a combination of the IID and the CCEP. Total calculated latency from start of IID to stimulation with the configuration of the example was 37ms. This latency is the summation of acquisition delay (7ms), 10ms delay added by smoothing (i.e. power at each point is the average of the 20 preceding samples) and the selected detection interval (20ms). A second example (Figure 3.E-G) shows IIDs detected with a bipolar montage in the amygdala. In this case a bipolar montage between LAT1 and LAT4 was used to reduce line noise contamination. Stimulation pulses were delivered to LAT2-LAT3. SPESs were sent with zero delay (Figure 3.E), 100 ms delay (Figure 3.F), or 200 ms delay (Figure 3.G). In the first case stimulation occurred during the IID spike, in the latter during the slow wave. For detailed performance analysis see section *CLoSES-RT Performance analysis*.

**Figure 3.**
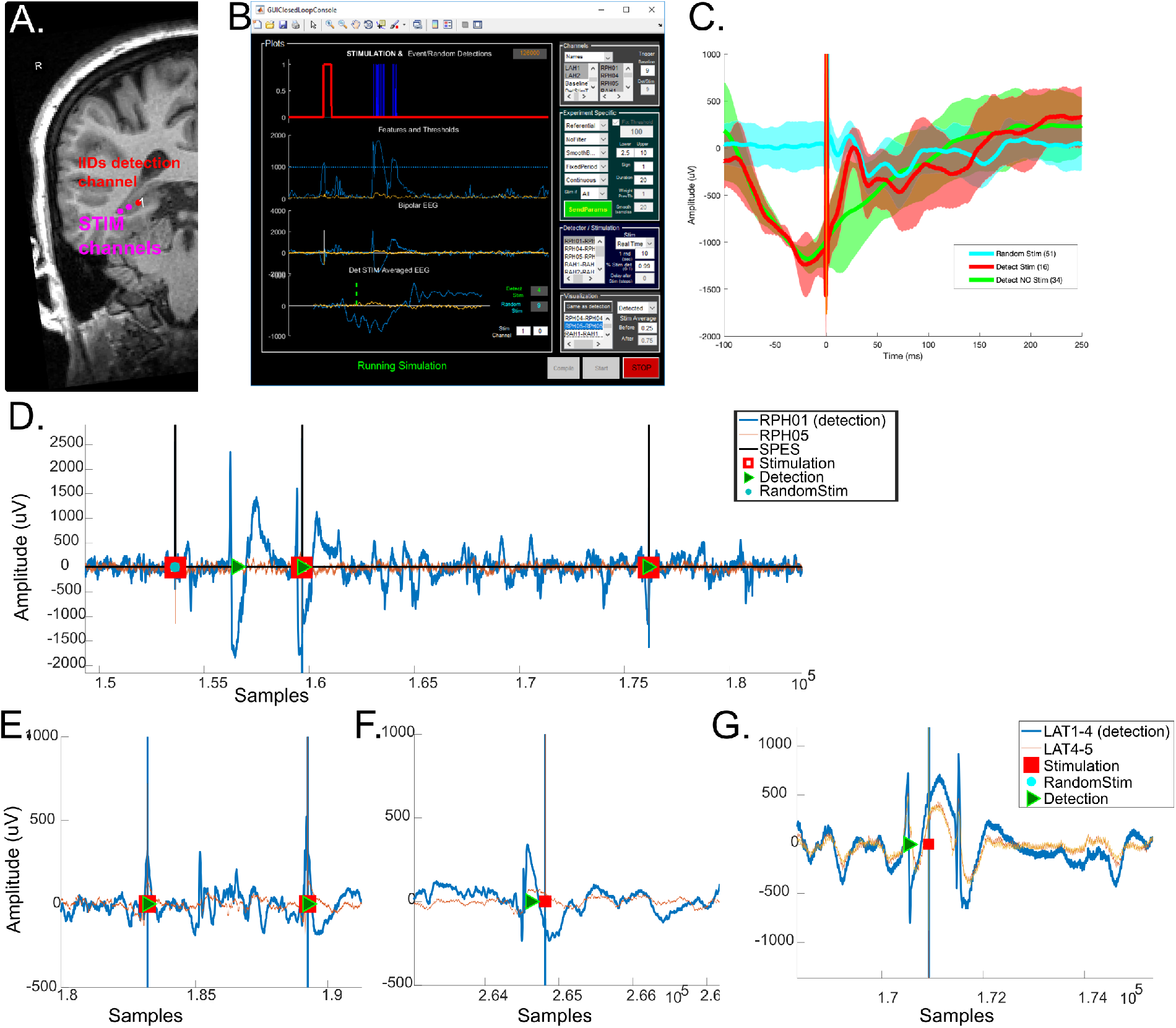
Examples of CLoSES-RT during closed-loop to IIDs experiments. A) IIDs were detected in the right hippocampus (red dot, RPH1), in the SOZ. When an IID was detected, single pulse stimulation was delivered to adjacent channels (magenta dots, RPH2-3). B) CLoSES-RT GUI during offline replay of IIDs detection in RPH1 using same data and configuration as in real-time experiment. First row: stimulation pulses (red) and detected events (blue). Second row: visualization of features, in this case power, and thresholds (magenta). Third row: raw iEEG. Last row: averaged iEEG locked to stimulation. Right panel allows configuration of parameters including real-time adjustments. The GUI and algorithms were the same for replay or real-time experiments. C) Averaged iEEG for random stimulation (cyan), detected stimulation (red) and events detected without stimulation. Before stimulation, there was a large amplitude decrease only when IIDs were detected regardless of stimulation. Following stimulation, detected and stimulated events were a combination of the response to stimulation and the IID. D) Example of iEEG recordings in two channels surrounding stimulation site in the same test as in A-C. E-G) Examples of different delays following stimulation in a different participant. E) zero delay – stimulation occured when an IID is detected. F) 100ms delay-stimulation pulse was sent 100ms after an IID is detected. G) 200ms delay – stimulation pulse was sent 200ms after an IID is detected. Note than in this case stimulation occurred during the slow wave. LAT: Left amygdala; RPH: Right posterior hippocampus channels; SPES single pulse electrical stimulation.

### 3.2 Application #2: CLoSES-RT detecting sleep spindles

We were interested in detecting sleep spindles and stimulating during the ongoing oscillation. Understanding the effect of SPES on these spontaneous events could have implications for sleep maintenance and memory consolidation. To this end, we selected neocortical channels with clear spindles on previous nights’ recordings. We configured CloSES-RT in the following way: band-pass filter between 11-15Hz (Butterworth order 2), smooth power (50ms window average), sleep spindle detection occurs if smooth power exceeded a threshold for 200-500ms. When a spindle was detected SPES stimulation was delivered in real-time. Referential or bipolar montages were selected for detection based on electrode location and 60Hz noise level. One to four channels were considered for detection. Figure 4 shows an example of detected and stimulated spindles, interleaved random stimulation, and detected but not stimulated spindles in the prefrontal cortex. Clear spindle activity can be observed in detected events regardless of stimulation, and clear CCEP after detected or random stimulation. In this example, total calculated latency from beginning of the spindle to stimulation with the configuration of the example was the sum of acquisition delay (7ms), filter’s group delay (81ms at central frequency, 13Hz), smoothing window (50ms), and detection interval (250ms) = 388ms. For detailed performance analysis see section *CLoSES-RT Performance analysis*.

**Figure 4.**
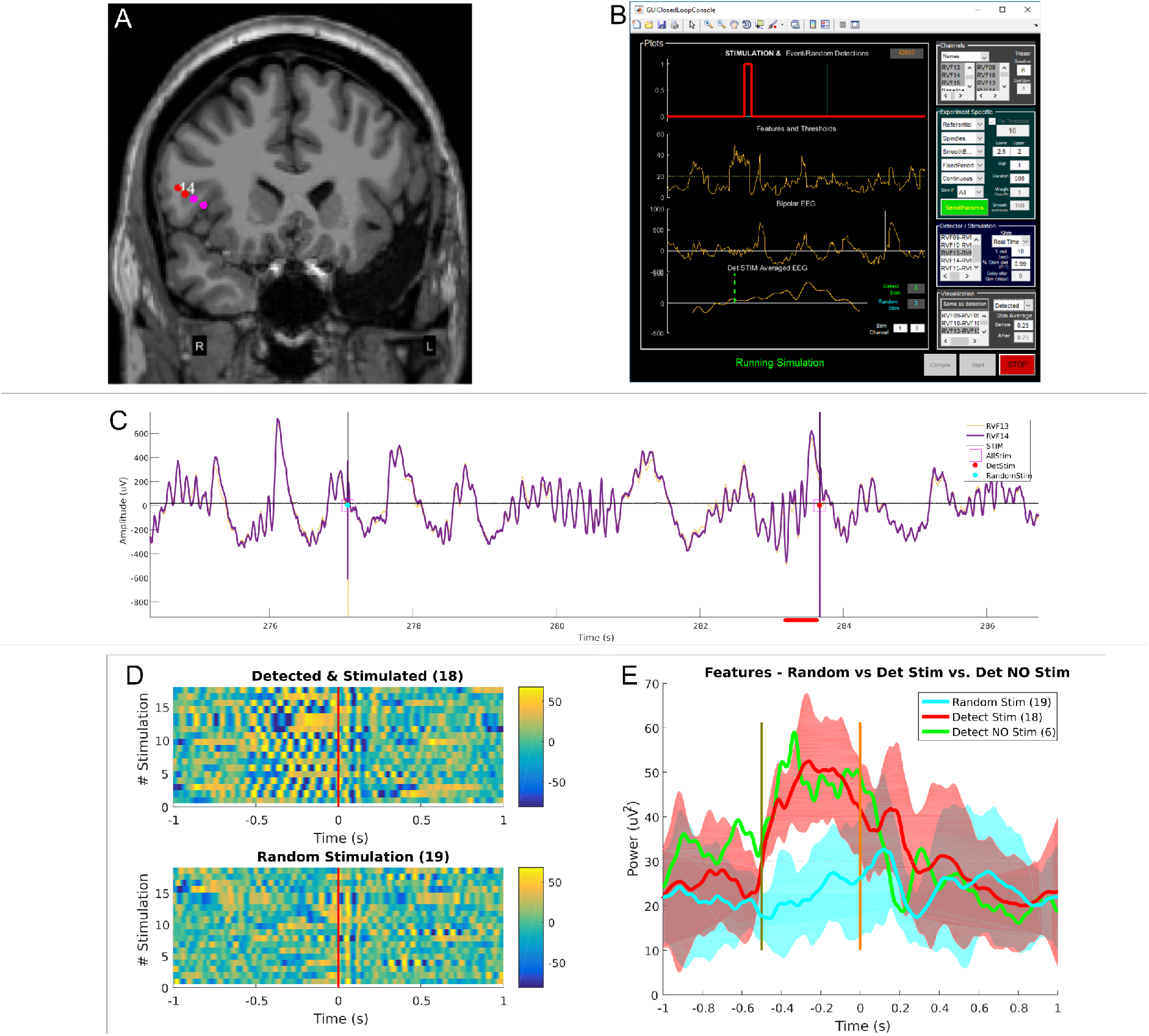
Example of CLoSES-RT during closed-loop to sleep spindles experiment. A) Spindles were detected in the Neocortex (red dots, RVF13 and RVF14) if power in any of these channels was above threshold for at least 250ms. When a spindle was detected, single pulse stimulation was delivered to adjacent channels (magenta dots, RVF). B) CLoSES GUI during offline replay of IIDs detection in RVF13 using same configuration as in real-time experiment. First row: detected and stimulated pulse (red) and random event (cyan). Note that the random pulse was within the refractory period and did not elicit stimulation. Second row: visualization of features, in this case power, and thresholds (magenta). Third row: raw iEEG. Last row: averaged iEEG locked to stimulation. Right panel allows configuration of parameters including real-time adjustments. The GUI and algorithms were the same for replay or real-time experiments. C) Example of iEEG recordings in two channels surrounding stimulation site. D) Per event stack plots of filtered iEEG. Top: Detected spindles followed by stimulation. Before stimulation, a spindle could be observed in the fil. Bottom: Random stimulation. Time zero (red line) indicates time of stimulation. E) Averaged features for random stimulation (cyan), detected stimulation (red) and events detected without stimulation (green). Time zero (red line) indicates time of stimulation, at -500ms, detection begins (green line). Following stimulation, detected and stimulated events were a combination of the response to stimulation and the IID. RVF: Right ventro-frontal.

### 3.3 CLoSES-RT performance and latency analysis

In CLoSES-RT, we specified a 1ms fixed step. Importantly, the tests required a low number of channels to detect events of interest at each fixed step. For 3-10 channels during IID detection each step takes less than 0.4ms (Figure 5.A-C) At each step iEEG was acquired, filtered, and power computed. For 6 channels, these steps took 0.13+/-0.001 ms for IIDs. Every 100ms data were sent back as UDP packets to the GUI for visualization and saved locally which took 0.14 +/-0.01ms. When detection occurred and SPES was sent took 0.24 +/-0.002ms (Figure 5.A-B).

**Figure 5.**
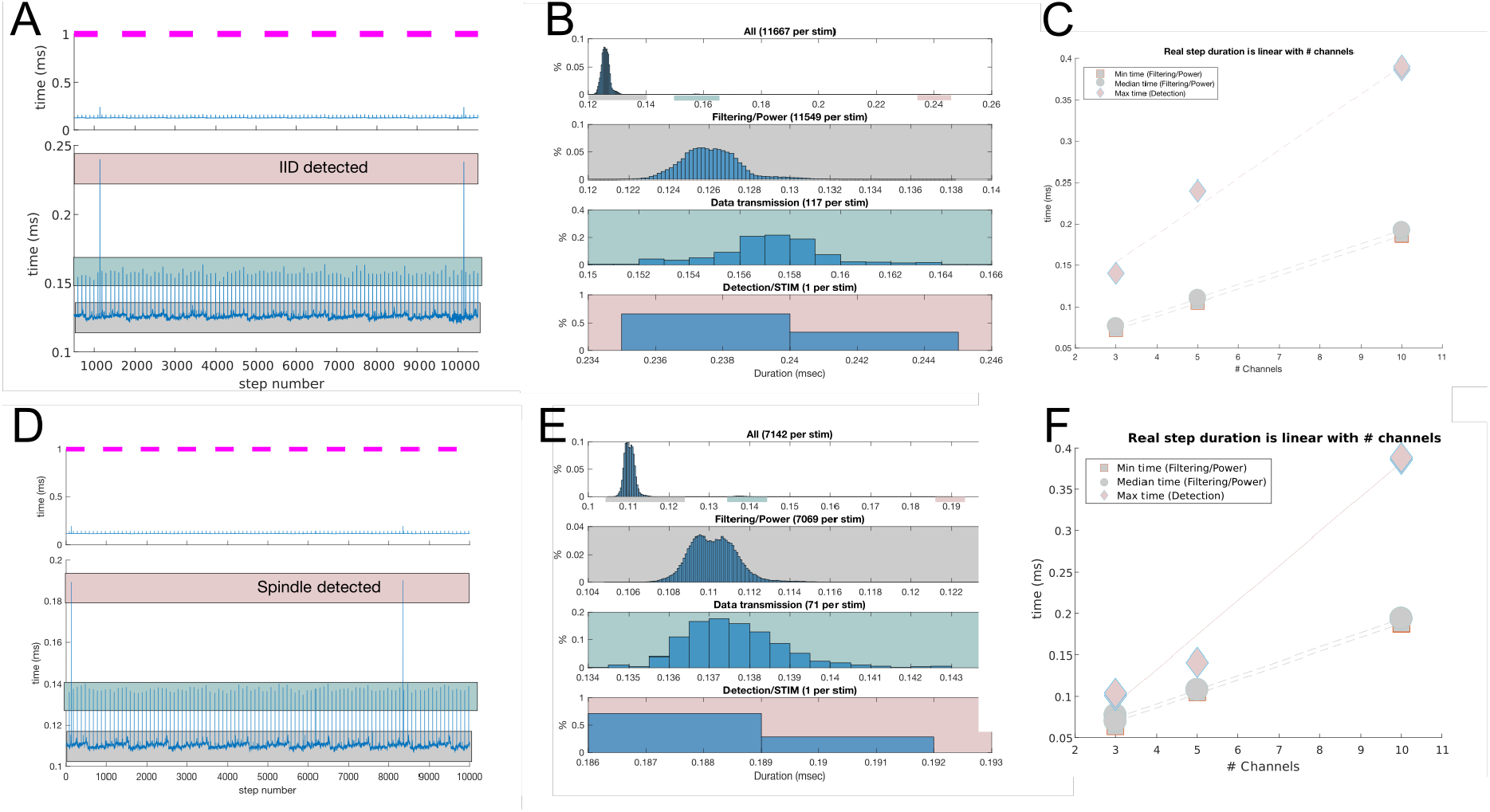
Time needed for each step when running the system in real-time on the EMU compatible rig for CLoSES-RT during closed-loop to IID and spindles experiments. Each step consists of iEEG acquisition at 2kHz, buffering, power computation, detection and sending of stimulation pulse. Fix step was set to 1ms (magenta lines) but it actually takes less than 0.25ms to perform these steps. A-B) For IIDs filtering and power calculation takes 0.126ms, when sending data back to GUI via UDP packets (every 100ms) it takes 0.14ms, when stimulation occurs it takes 0.24ms. C) The real duration of a step linearly increases with number of channels. Min and median duration correspond to filtering and feature calculation steps. D-E) For spindles only filtering takes 0.11ms, when sending data back to GUI it takes 0.14ms, when stimulation occurs it takes 0.19ms. F) The real duration of a step linearly increases with number of channels. Max corresponds to steps when an IID was detected and stimulation pulse sent. All situations were well below 1ms.

Similarly, for spindles for 3-10 channels during IID detection each step took less than 0.4 ms (Figure 5.D-F). As an example, for 3 channels, acquisition, filtering and feature calculation took 0.11+/-0.001ms. When data were sent back to host for visualization, took 0.14 +/-0.001ms. When SPES was delivered, took 0.19 +/-0.002ms (Figure 5.D-E).

In both tests real step duration was much less than the one millisecond fixed step. The real duration of a step increased linearly with number of channels. Even with 10 channels the real duration was well below 1ms (Figure 5.C and 5.F).

### 3.4 Application #3: CLoSES-SEM during cognitive flexibility task

CLoSES-SEM was used during a Multi-Source Interference Task (MSIT) to study the effect of stimulation on cognitive flexibility. An encoder/decoder model based on multi-band log power input features was used to estimate hidden cognitive state (Yousefi et al., 2019a). Details on MSIT and corresponding model can be found elsewhere (Basu et al., *submitted*). During MSIT CLoSES-SEM was configured in the following way: Theta, Alpha, and High Gamma multi-band filter of pre-determined bipolar channels were implemented; aggregate logarithmic spectral power in the three frequency bands were calculated over a time window of 2 seconds following image onset; a neural decoder model was used to estimate a hidden flexibility state using the calculated log power; if estimated mean state value exceeded a pre-determined threshold a short train of 130 Hz stimulation was triggered on the following trial (following image onset). As a safety measure, only 5 out of 10 trials could receive stimulation within a sliding moving window including 10 trials.

To obtain a participant specific encoder/decoder model, each participant performed at least 128 trials of MSIT without stimulation and open-loop stimulation while their behavior and iEEG were simultaneously recorded. Simulation datasets with all channels were created from these trials and ran offline on replay with CLoSES GUI. This allowed determination of a reduced set of spectral features and a decoder model tailored for each participant (Yousefi et al., 2019a). This dataset was also used to estimate an initial threshold for stimulation. During the closed-loop tests, CLoSES-SEM was loaded with the optimized participant specific model and configuration file. There was a block (64-96 trials) without stimulation, followed by 1-2 blocks (64-128 trials) with CLoSES-SEM, followed by one more block without stimulation. Figure 6 shows an example of CLoSES-SEM during MSIT.

**Figure 6.**
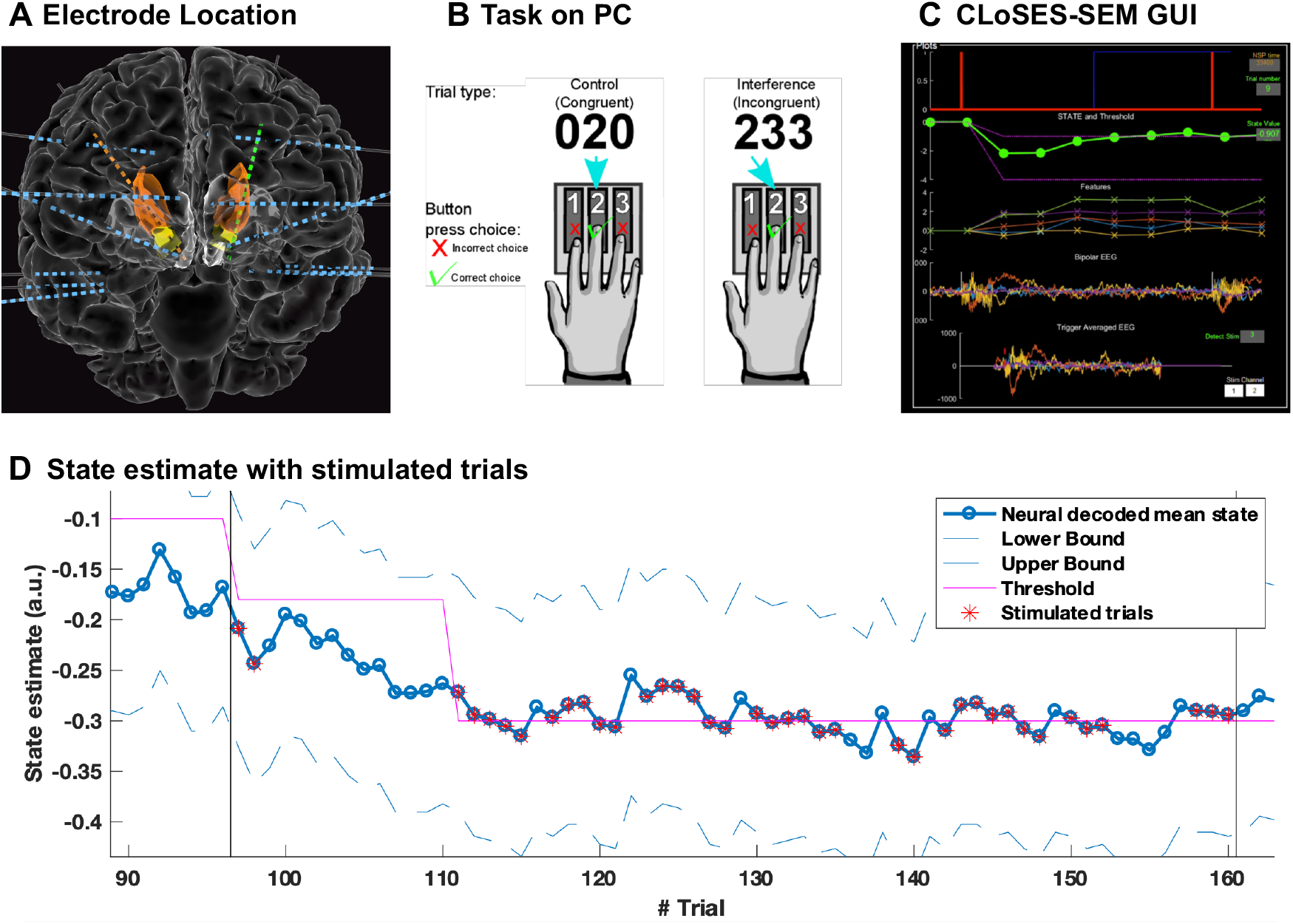
Example of CLoSES-SEM during closed-loop MSIT experiment. A) In this participant, power in Theta, Alpha and High-Gamma bands in several regions were used as input to the state estimate model. Stimulation was in dACC if mean state estimate was above the higher threshold. B) Illustration of MSIT task. C) CLoSES-SEM during replay of this participant’s data. D) State estimate based on multiband power. Up to trial 96-no stimulation, Trials 97-161 closed-loop experiments with stimulation on following trial, next trials: no stimulation. Threshold is shown in magenta.

### 3.5 Application #4: CLoSES-SEM during emotion regulation task

CLoSES-SEM was used during an Emotion Conflict Resolution (ECR) task to study the effect of bstimulation on emotion regulation. An encoder/decoder model based on theta band coherence input features across regions was used to estimate hidden state. Details on the implementation of ECR can be found elsewhere (Paulk et al., *submitted*). During ECR CLoSES-SEM was configured on the following way: Theta band filter, coherence, detection if boundary of state estimates were above/below respective thresholds, and stimulation on the following trial (following image onset). As a safety measure, only 5 out of 10 trials could receive stimulation.

Similar to MSIT, to obtain a participant specific encoder/decoder model, each participant performed at least 128 trials of ECR without stimulation while their behavior and iEEG were simultaneously recorded. Simulation datasets with all channels were created from these trials and offline ran on replay with CLoSES GUI to create participant specific models. Given the large number of possible coherence pairs, replaying these datasets was an important step to reduce feature dimensionality. During the closed-loop tests, CLoSES-SEM was loaded with the optimized participant specific model and configuration file. There was a block (64 trials) without stimulation, followed by 1-3 blocks with CLoSES-SEM, followed by one more block without stimulation. Figure 7 shows an example of CLoSES-SEM during ECR test in which bi-directional control was achieved (i.e. when state estimate decoded from neural features was above threshold stimulate on one site, when it was below another threshold stimulate on a different location).

**Figure 7.**
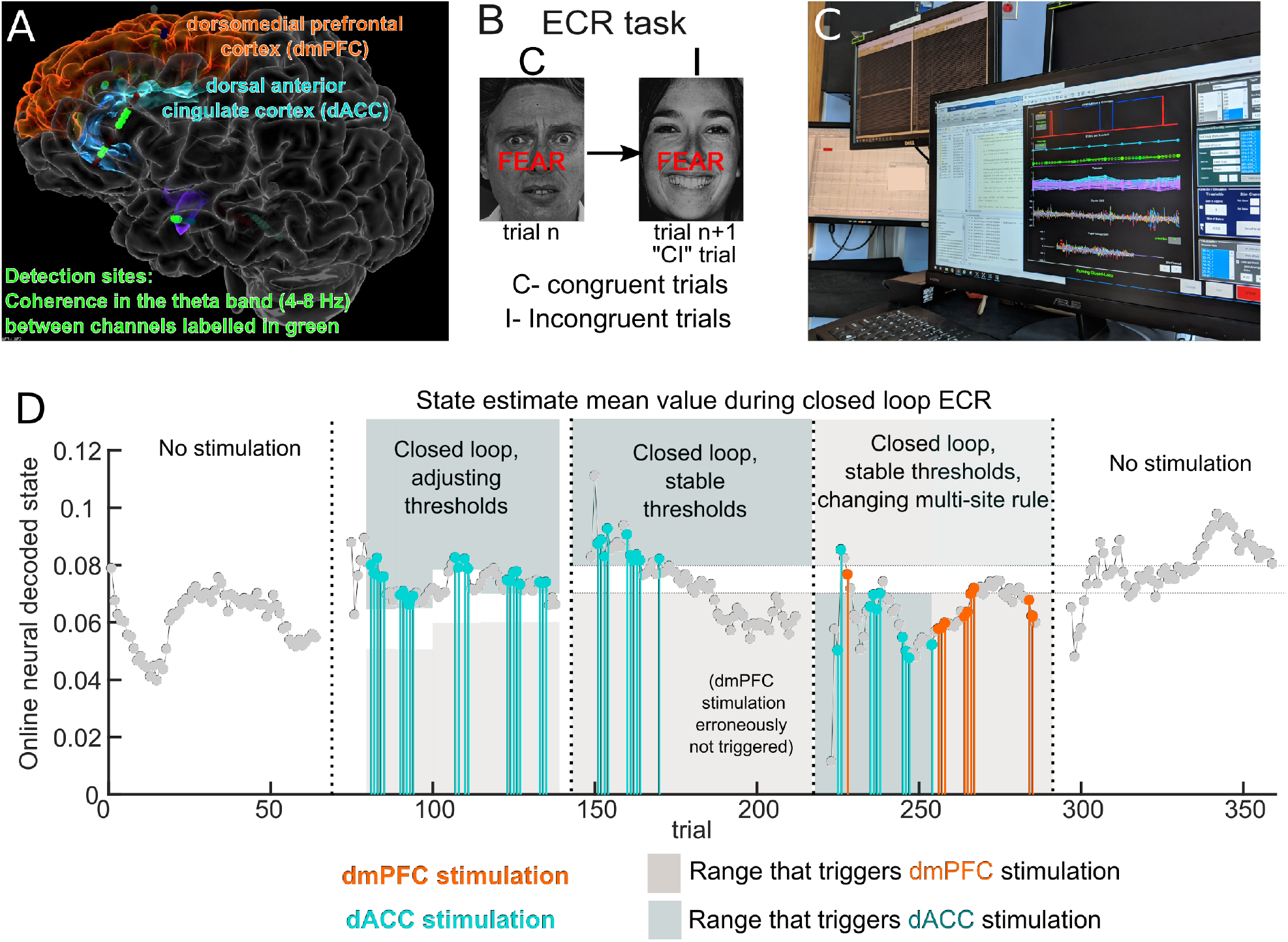
Example of CLoSES-SEM during closed-loop ECR experiment. Bi-directional control was achieved. A) Illustration of ECR task. B) In this participant, coherence in Theta band between several regions (in green) were used as input to the state estimate model. Stimulation was in dmPFC if lower bound of state estimate was below the lower threshold and in dACC if upper bound of state estimate was above the higher threshold. C) Five blocks of ECR, 1-no stimulation, 2-4 closed-loop experiments with stimulation on following trial, 5-no stimulation. Note in block 3 how the mean state gets lower with subsequent stimulations. dmPFC: dorso-medial prefrontal cortex; dACC: dorsal anterior cingulate cortex.

### 3.6 CLoSES-SEM performance and latency analysis

On CLoSES-SEM, the continuous steps (acquisition, filtering and feature calculation) must occur within the fixed step duration of 10 ms, but we could relax this constraint for blocks that are executed only once per trial (e.g. decoder model). The real duration of a step increased linearly with the number of channels for filtering, and feature (power and coherence) calculation (Figure 8.A and 8.D). For logPower up to 50 channels and coherence up to 20 channels the real duration of the step was below 10ms. For the one time per trial when decoder model ran, duration was linear with number of features (Figure 8.A and 8.D). As an example, for MSIT, 50 channels, each filtered in three frequency bands, and power calculated on 150 filtered signals, then averaged during one epoch, resulted in 150 input features to the decoder model. The actual duration for each step was (mean+/-std): when only filtering: 0.53+/-0.005 ms; when computing log power: 0.68+/-0.005 ms; the one step per trial when decoder model ran: 54+/-0.1 ms; in the following step rate transitions were adjusted: 0.71+/-0.02; then detection occurs: 1.21+/-0.009 ms; and trial by trial information was saved: 0.54+/-0.01 ms (Figure 8.B &C). As an example, for ECR 20 channels filtered in the theta band, followed by coherence on all 190 pairs during 2 epochs, resulted in 374 603 input features (six features were not used as inputs in this example). Actual duration for each step 604 was (mean+/-std): when only filtering: 0.25+/-0.002ms; when coherence was computed: 4.92+/-605 0.004 ms; the step when decoder model ran: 105.8+/-0.03 ms; in the following step rate transitions 606 were adjusted: 0.315+/-0.004; then detection occurred: 0.91+/-0.004 ms; and trial by trial 607 information was saved: 0.59+/-0.008 ms (Figure 8.D, E &F). Thus, all continuous computations were well within the 10ms fixed step. It is unlikely that more than 200 pairs of coherence would be required, but if more channels were of interest the fixed step could be adjusted. As shown in the examples, Decoder model step was dependent on number of features (54ms or 6 steps for 150 features, 105ms or 11steps for 374 features).

**Figure 8.**
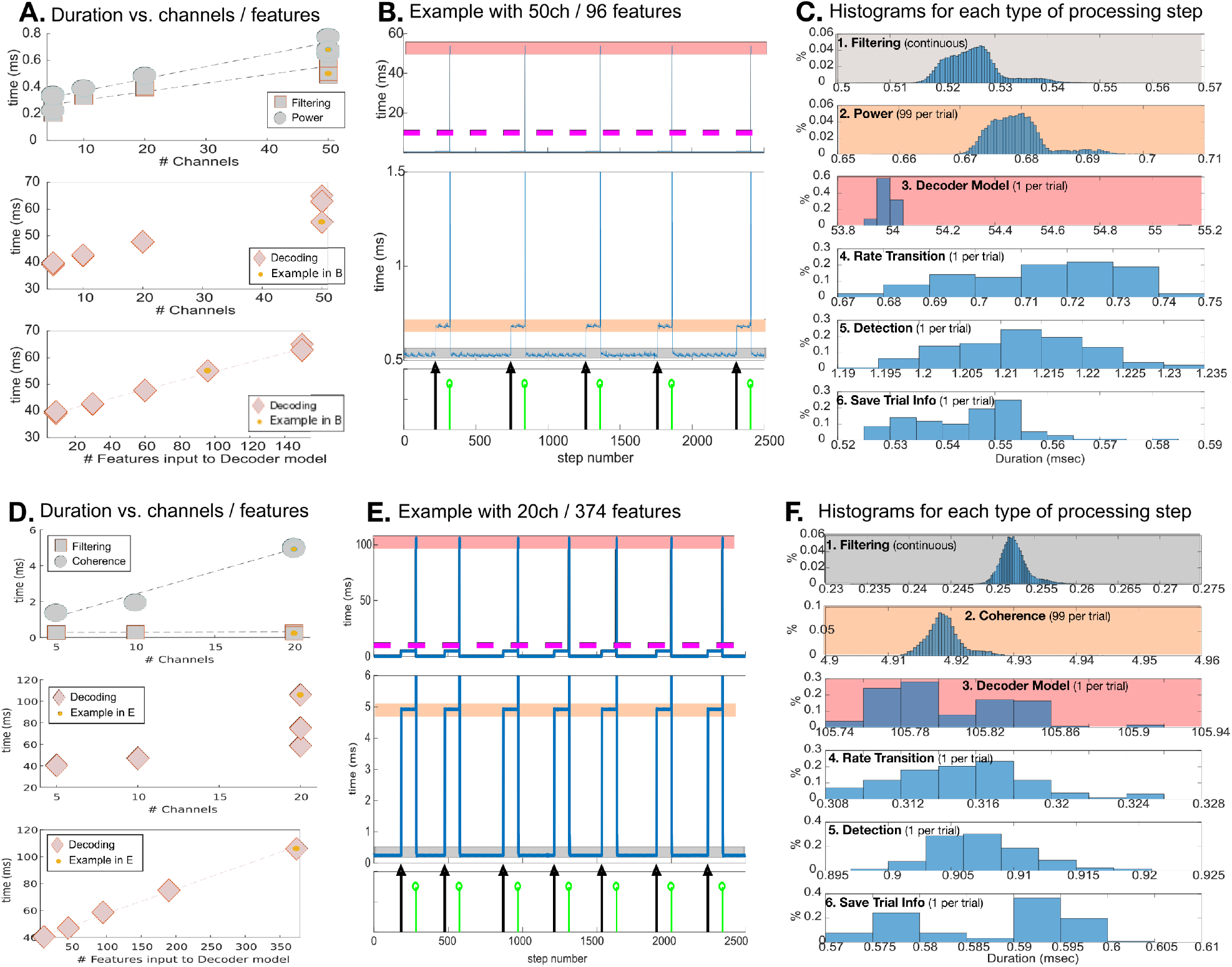
Time needed for each step when running CLoSES-SEM in real-time on the EMU compatible rig. A: During MSIT tests (Log Power calculation, stim at following trial) the duration of each step increased linearly with the number of channels for filtering (circles) and power calculation (squares). The duration increased linearly with number of features for decoder model estimation (diamonds). B: Example of the time that took processing each step during MSIT for 50 channels, 96 features (marked with a yellow dot in A). Top part shows example of five consecutive trials, middle part a zoom for 0.5-1.5ms. Black arrow indicates time of image onset (beginning of trial). Green arrow indicates time when continuous power was averaged, and decoder model ran. Magenta line indicates step fix duration (10ms). C: Histograms of time that each step took dividing by type of processing step for the same example of B. Histograms are ordered by when the type of steps was performed per trial. 1-Filtering is performed at every step (“continuously”) and took 0.53+/-0.005 ms per step. 2-LogPower was computed from image onset to decoder time and took 0.68+/-0.005 ms. 3-Estimating the state with the decoder model took 54+/-0.1 ms, which is longer than the 10ms fix step. Importantly, the decoder step is non-critical as it was run only once per trial (during one step) and the system had ∼1 second to recover before the following trial. 4-A rate transition must run after the decoder step finished, to compensate for the extra allotted time (0.71+/-0.017 ms). 5-Detection based on the state estimated value took 1.21+/-0.01 ms. 6-Saving trial information took 0.55+/-0.01 ms. Y axis indicates percentage of steps for each duration. Gray, orange and pink boxes in B and C relate the histograms to the continuous plots. Step was set to 10ms, meaning that the next processing step starts every 10ms. D: During ECR tests (Coherence calculation, stim at following trial) the duration of each step increased linearly with the number of channels for filtering (circles) and power calculation (squares). The duration increased linearly with number of features for decoder model estimation (diamonds). E: Example of the time that took processing each step during ECR for 20 channels, 190 coherence pairs, averaged over 2 epochs, 374 decoder input features (marked with a yellow dot in D). Top part shows example of seven consecutive trials, middle part a zoom for 0-6ms. Black arrow indicates time of image onset (beginning of trial). Green arrow indicates time when continuous power was averaged, and decoder model ran. Magenta line indicates step fix duration (10ms) F: Histograms of time that each step took dividing by type of processing step for the same example of E. Histograms are ordered by when the type of steps was performed per trial. 1-Filtering is performed at every step (“continuously”) and took 0.25+/-0.002 ms per step. 2-Coherence was computed on 190 pairs from image onset to decoder time (4.92+/-0.004 ms per step). 3-Estimating the state with the decoder model took longer than the fix step, 105.8+/-0.03 ms. Importantly, the decoder step is non-critical as it was run only once per trial (during one step) and the system had ∼1 second to recover before the following trial. 4-A rate transition must run after the decoder step finished, to compensate for the extra allotted time (0.315+/-0.004 ms). 5-Detection based on the state estimated value took 0.91+/-0.004 ms. 6-Saving trial information took 0.59+/-0.008ms. Y axis indicates percentage of steps for each duration. Gray, orange and pink boxes in B and C relate the histograms to the continuous plots. Step was set to 10ms, meaning that the next processing step starts every 10ms.

To push the system outside normal operation limits, we tested ECR with 325 coherence pairs (all combinations from 26 channels) and found that coherence took slightly longer than 10ms. Even though execution took sometimes longer than the fixed step, because the rate transitions and because coherence was only computed during certain epochs of a trial, CLoSES could still execute 616 successfully. Indeed, CLoSES-SEM continued running successfully for 4 hours (876 trials). As a comparison, during closed-loop tests we ran 192 trials. Importantly, if larger number of features are required, the step size of 10ms could be increased, effectively allowing for any number of channels and features.

## 4 Discussion

In this paper, we have presented CLoSES: a research tool that flexibly solves several challenges in closed-loop stimulation with iEEG in humans. We demonstrated the use of our closed-loop stimulation system in participants with implanted electrodes in the EMU in a variety of paradigms: 623 1) in real-time detection of spontaneous events or trial by trial during cognitive tasks; 2) with very 624 low latency to stimulate during fast events like IIDs or with longer detection duration for slow 625 neural rythms; 3) triggering stimulation with uni or bi-directional control at specific times; 4) 626 during different brain states, such as sleep, awake and rest, and during the performance of cognitive 627 tasks. We reported latency for different situations and tested the limits of the system both for replay 628 and real-time operation. CLoSES latency is lower than most BCI application (Wilson et al., 2010).

Brain oscillations at particular frequency bands are related to pathology (e.g. high-frequency oscillations in epilepsy, Beta oscillations in Parkinson) and physiology (e.g. Mu rhythm in 631 movement, Gamma in attention, Ripples in memory, spindles during sleep). Duration, amplitude, location, interaction, and underlying mechanisms are different for different neuronal events. Our flexible closed-loop framework can detect these different oscillations and allows probing rhythms 634 and cross-frequency interactions. CLoSES successfully produced responsive stimulation during short latency transient pathological IIDs, sleep spindles, theta band, or high gamma activity. By detecting transient or oscillations based on its particular characteristics, incorporating problem-specific control algorithms, and stimulating at precise times our system could help answer a variety 638 of pressing neuroscience questions (Lozano et al., 2019; Widge and Miller, 2019).

### 4.1 Comparison to Existing Systems

In recent years, tremendous advances in software and hardware for closed-loop tests have been 641 made. Development of flexible tools, freely available to the community, is important for neuroscience and clinical research, allowing the field to grow. Perhaps a paradigmatic example, Open Ephys is comprised of low cost acquisition equipment and a GUI (Siegle et al., 2017) that is open-source, and allows researchers to add their own modules. The Open Ephys GUI is written in 645 C++ and connects to the acquisition board through USB. It is mainly targeted to neuroscience basic research in animals but has also been used to record scalp EEG and EMG activity with inexpensive hardware (Black et al., 2017). BCI2000 (Schalk et al., 2004) has been widelly used by the BCI community. It is also written in C++ and it’s newest implementation allows web based control 649 (Milsap et al., 2019). Real-Time eXperiment Interface (RTXI), provides a modified real-time Linux OS and was written in C++ (Patel et al., 2017). RTXI is also aimed at basic science research. Similarly, Falcon was also written in C++ and provides sub millisecond latencies, making it ideal for closed-loop tests recording from population bursts (Ciliberti and Kloosterman, 2017). Instead 653 we focused on LFP acquisition, computing local and network features. Using UDP or direct boards for acquisition and fixed steps provided by Simulink Real-Time, result in short controlled latencies. Moreover, as CLoSES was developed with Simulink and MATLAB, it is easy to add new functionalities and visualization capabilities. In this way, we expect to reach a large number of clinical and translational researchers even if they have limited programming skills.

CLoSES is unique as it is targeted to be used in the hospital room with existing, robust multichannel acquisition systems and is flexible in how it can be utilized from the features chosen for detection through to the outputs that can be delivered to control stimulation or other actuators. Thus, we believe that CLoSES bridges a challenging gap, effectively complementing existing 662 systems, but extending the strength of the closed loop approach in a hospital setting.

### 4.2 Limitations and Future Work

It could be argued that a limitation of our system is that MATLAB and Simulink are paid software. 665 However, we believe that the benefit of using a modular intuitive programing platform such as Simulink and the extensive visualization possibilities provided by MATLAB outweighs the cost. Moreover, the widespread use of MATLAB in the scientific community makes it easy for other groups to develop their own paradigms based on the CLoSES platform.

We foresee several technical additions in the near future. Stimulation artifact could obscure recordings. Initially, we implemented notch filters, but they were too slow for most applications. In practice, we avoid stimulation artifact effects on detection, by setting a refractory period following stimulation. Incorporating new artifact-removal techniques (Zhou et al., 2018) could 673 allow detections closer to stimulation and newer amplifier systems are increasingly designed to 674 decrease the stimulation artifact in general.

Only a few of the most common features were implemented. Given the modular approach of our platform, new features could be easily added. For example, in the case of neural oscillations, 677 stimulating at a particular phase could have different effects (Widge et al., 2018a). Therefore, it would be interesting to incorporate phase-locked stimulation following detection of oscillations. Finally, another path for development is the integration of the current system with field programmable gate arrays (FPGAs). FPGAs have been applied to large throughput systems (e.g. 681 10,000 input channels and deep learning recurrent networks) for real-time neural decoding (Heelan et al., 2018).

### 4.3 Future application

#### Precision medicine and adaptive DBS

Therapeutically, CLoSES could be used as prototyping 685 platform in the EMU before implanting a closed-loop DBS (Widge et al., 2017). The pipeline could be the following: 1) Tentative detection and control targets are obtained from neuropsychological testing, questionnaires and imaging; 2) Electrodes are implanted in these putative 688 target areas; 3) Run tasks while stimulating with CLoSES to precisely identifying effective regions and better understand the effect of stimulation in each patient; 4) Implant chronic DBS system in the effective regions.

#### Study of human physiological and pathological neural mechanisms

As a research tool, 692 CLoSES streamlines the testing of a wide variety of questions in humans. CLoSES-RT has been used to investigate the mechanisms and roles of sleep spindles and epileptic IIDs. CLoSES-SEM has been used to investigate the effect of stimulation during cognitive flexibility and emotion regulation tasks. Many other rhythms and cognitive questions can be effectively probed using the 696 closed-loop approach made more turn-key through CLoSES.

#### CLoSES in other clinical environments

CLoSES was developed and tested for use in the EMU, but could be utilized in other environments where we record electrophysiological signals and perform stimulation. In particular, in the operating room (OR), we could perform acute closed-700 loop tests with novel electrodes to study the response at different scales and study the response of the brain during diverse brain states, such as wake and general anesthesia. CLoSES is currently ready to be utilized in the OR. In outpatient clinics, we could easily adapt the input (acquisition) and output modules to adapt CLoSES to closed-loop tests using TMS-EEG or TMS-MEG. Combining results of closed-loop paradigms for non-invasive and direct stimulation, related to 705 physiological and pathological activity, at different scales, and during different brain states, a more complete understanding of brain mechanisms and the effect of stimulation could be achieved.

With the release of the CLoSES platform as open-source software simultaneously with this paper publication, we hope that a wider number of research questions could be answered, and a large 709 number of therapeutic clinical biomarkers could be evaluated.

## 5 Conclusions

CLoSES successfully triggers stimulation based on real-time detection and decoding of brain signals, during cognitive tasks, awake rest, and sleep. CLoSES provides a flexible platform to 712 implement a variety of closed-loop experimental paradigms in humans including: 1) probe oscillations and understand their mechanisms, role in information transfer, and state maintenance; 2) study possible modulatory effect of physiological and pathological electrophysiological signals on the brain’s response to stimulation; 3) identify and evaluate potential electrographic biomarkers 716 of neurological and psychiatric disorders; 4) and test patient specific stimulation targets and control signals before implanting a therapeutic device. We expect that studying the effect of stimulation on brain dynamics and the modulation of brain states on stimulation response will lead to important insights into the mechanisms of normal and pathological brain activity.

## Data Availability

CLoSES code and example data is available on GitHub.

https://github.com/Center-For-Neurotechnology/CLoSES-SEM

https://github.com/Center-For-Neurotechnology/CLoSES-RT

## 6 Acknowledgments

This work was supported in part by the Tiny Blue Dot foundation, NIH grant K24-NS088568, NIH grant R01-NS062092, NIH grant R01NS079533 (WT), and the Defense Advanced Research 722 Projects Agency (DARPA) under Cooperative Agreement Number W911NF-14-2-0045, issued 723 by the Army Research Office contracting office in support of DARPA’S SUBNETS program. RZ 724 was supported in part by the MGH ECOR Fund for Medical Discovery Postdoctoral Fellowship 725 Award. The views, opinions, and/or findings expressed are those of the authors and should not be 726 interpreted as representing the official views or policies of the Department of Defense or the U.S. 727 Government.

